# Development of a colorimetric loop-mediated isothermal amplification diagnostic assay for deer tick virus

**DOI:** 10.1101/2020.08.24.20181248

**Authors:** Brandon G. Roy, Morgan A. Decker, Lauren Vickery, Eric J. Ryndock

## Abstract

Deer tick virus (DTV) is an emerging pathogen in North America. This virus can cause nervous system complications such as encephalitis in humans. Further, no data has been surmounted around long-term effects of infection from DTV patients across variable age groups. Diagnostic tools of DTV used by government laboratories are based on RT-PCR using patient serum or ticks. This paper explores the feasibility of a colorimetric loop-mediated isothermal amplification (LAMP) assay to create a point-of-care diagnostic methodology for use in field and in primary care. LAMP consists of six primers that bind to target DNA and amplifies variable length nucleotide strands that can be visualized through side reactions or via electrophoresis. First, a viable LAMP primer set, and a primer set that dimerizes and amplifies DNA regardless of compatibility were created in silico and validated in vitro. Then, a specific LAMP assay was developed. Our findings showed this method can be performed within 30 minutes and can measure with limits of detection comparable to PCR.

## Introduction

Deer tick virus (DTV) has a positive-sense, single-stranded RNA genome and belongs to the genus *Flavivirus* in the family *Flaviviridae*. Other members of this virus genus and family are dengue virus, Zika virus and West Nile virus. The genome length of DTV is roughly 10,800 bases and it known to cause tick-bourne encephalitis (Artsob et al. 2001).

It is common that members of *Flaviviridae* are vectored through arthropods such as mosquitoes and ticks. The increasing presence of DTV throughout North America is coupled with various tick species (CDC 2019). DTV is also referred to as the second lineage Powassan virus (POW2), deviating from a native strain in Europe and Russia. POW1, or Powassan virus lineage I, has a variable genetic diversity along the theoretical divergence cutoff at 84% nucleotide identity to DTV, 94% amino acid identity, and is estimated to have diverged around 200 years ago (Pesko et al. 2010). This cutoff provided the difference between the two Powassan viruses on the cladogram of *Flaviviridae* (Telford III 1997). Patient testing for DTV is usually performed in response to severe cases where symptoms arrive suddenly. Most people tested for DTV have symptoms of acute encephalitis or overactive inflammation of the brain, which leads to death in some cases without proper medical treatment (Tavakoli et al. 2009; Solomon et al. 2018). Remaining cases go unbeknownst because testing measures are not approached from typical or asymptomatic standpoints as other tick-borne infections. Current diagnostic testing is limited to the Center of Disease Control and its partnering laboratories using either reverse transcription-polymerase chain reaction (RT-PCR) or immunohistochemical techniques (Morozova et al. 2002; CDC 2019). A sample of serum or cerebral spinal fluid is collected and sent to one of the offsite diagnostic centers, further complicating the testing process and taking up to a few days for results to become available.

A more time effective detection method than RT-PCR would be beneficial for point-of-care testing (POCT), as well as for field surveys of tick samples that are performed by state game commissions. Three different species of ticks, i.e., *Ixodes cookie, Ixodes marxi*, and *Ixodes scapularis*, have the potential to transmit DTV from their main vertebrate hosts, i.e., woodchucks, squirrels, and white-footed mice, to humans (Brackney et al. 2010). The white-tailed deer, *Odocoileus virginianus*, is also a known host to *Ixodes scapularis*. DTV has no known pathology on these intermediate mammal hosts, but there is increasing concern towards human infection occurrence (CDC, 2019). The main tick vector that raises human concern is *Ixodes scapularis*. This tick species has a parasitic relationship with white-tailed deer and other species of deer that humans encounter frequently in rural areas (Brackney et al. 2010). This host relationship is unique to DTV and unlike the previously mentioned POW1 strain. The prevalence of outdoor recreation and leisure activities give exposure to the tick vectors and potential pathogens to be transmitted.

Monitoring the presence of DTV provides the first steps to surveying disease spread and to developing mitigation efforts. A rapid DTV detection assay like colorimetric loop-mediated isothermal amplification (LAMP) would prove useful for field surveys and POCT. LAMP is an alternative nucleic acid amplification technique that has gained popularity in the past decade (Tomlinson 2008; Luo et al. 2011; Chen et al. 2014; Lau et al. 2015; Lamb et al. 2018; Shin et al. 2018; Anupama et al. 2019). LAMP is a nucleotide amplification process that occurs through the binding of specially designed primers at approximately 60°C that generates variable ladder structures that can be visualized in two manners. Colorimetric LAMP does not require expensive and sophisticated equipment or visualization techniques other than the naked eye. Additional variants of LAMP assays, such as RT-LAMP, have surmounted for the specific detection of RNA as opposed to cDNA which was utilized in this report (Chen et al. 2014). In addition, LAMP is inexpensive and quick. For example, the Colorimetric Warmstart® MasterMix (New England Biolabs, Cat# M1800S) LAMP technique, utilized in this paper, is up to five times quicker than PCR and changes color for visual assessment of the results (Nagamine et al. 2002). The colorimetric test starts out as pink and turns a bright yellow color if the result is positive. Color change is dependent on the release of protons through nucleotide assimilation on target DNA (Mitra et al. 2019). The amplified DNA can then be confirmed through electrophoresis on an agarose gel followed by staining with ethidium bromide or Coomassie Blue® to display banding, validating the presence of DTV in the tick sample tested. Additional efforts have been made available to educate and summarize LAMP technology in recent literature (Becherer et al. 2020; Roy et al. 2020). In this paper we present our preliminary data on the development of a colorimetric LAMP detection of DTV and discuss perspectives of our research for POCT.

## Materials and Methods

### Nucleotide sequence analysis and homology analysis for DTV strains

The greatest amount of genetic variance between strains of DTV were estimated through a maximum composite likelihood analysis method. The greatest amount of conservation was determined to be at the RNA-dependent RNA Polymerase (RdRp) coding site using the codon based Z-test. Two bioinformatics methods, i.e., the codon-based Z-test, and the Neighbor-joining phylogenetic tree method, were used to analyze the RdRp coding region or full DTV genome to determine nucleotide sequence conservation. Phylogenetic analyses were performed using the software suite MEGA7: Molecular Evolutionary Genetic Analysis version 7.0 (Kumar et al., 2015) using 22 DTV accessions retrieved from GenBank, an open-source genetic sequence database hosted by the National Institute of Health.

### Codon based Z-Test

Alignment tools were utilized to investigate the homology between the conserved sequences of DTV. Positive selection for the resistance to mutation was analyzed through the codon-based Z-test as defined by Equation 1. When analyzing the homology, the null hypothesis is regarded such that the sequences are not similar and only by rejecting the null hypothesis when *p* < 0.05, can we assume that the sequences are considered homologous. The analysis used four different sequences found in the United States with variable accession ID prefixes to provide additional variability to the analysis.

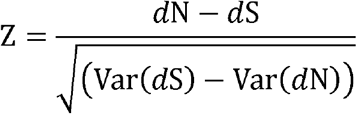

**Equation 1. The Z-test for codon homology testing across multiple DTV genomes**. Variables denoted in the equation are defined as the number of nonsynonymous substitutions per nonsynonymous site (dN), number of synonymous substitutions per synonymous site (dS), and the respective variances of these variables (Var(dN) and Var(dS)).

### Primer design

Primers for the LAMP assay were designed through the PrimerExplorerV5 LAMP Primer Designer using the following criteria: (1) genomic regions with a GC ratio of 50-60%, (2) no more than four successive G’s or C’s to prevent primers from binding too tightly to the target DNA strand, and (3) stability of F3 and B3 primer ends be lower than −4.00 to create the most stable 5’ and 3’ loop structures for LAMP proliferation (Wang et al. 2015). The region of the viral genome that was selected for primer design was NS5a that codes the non-structural polymerase.

### Primer constructs

Two primer constructs are evaluated in this paper (Table 1 and Table 2).

**Table 1.**
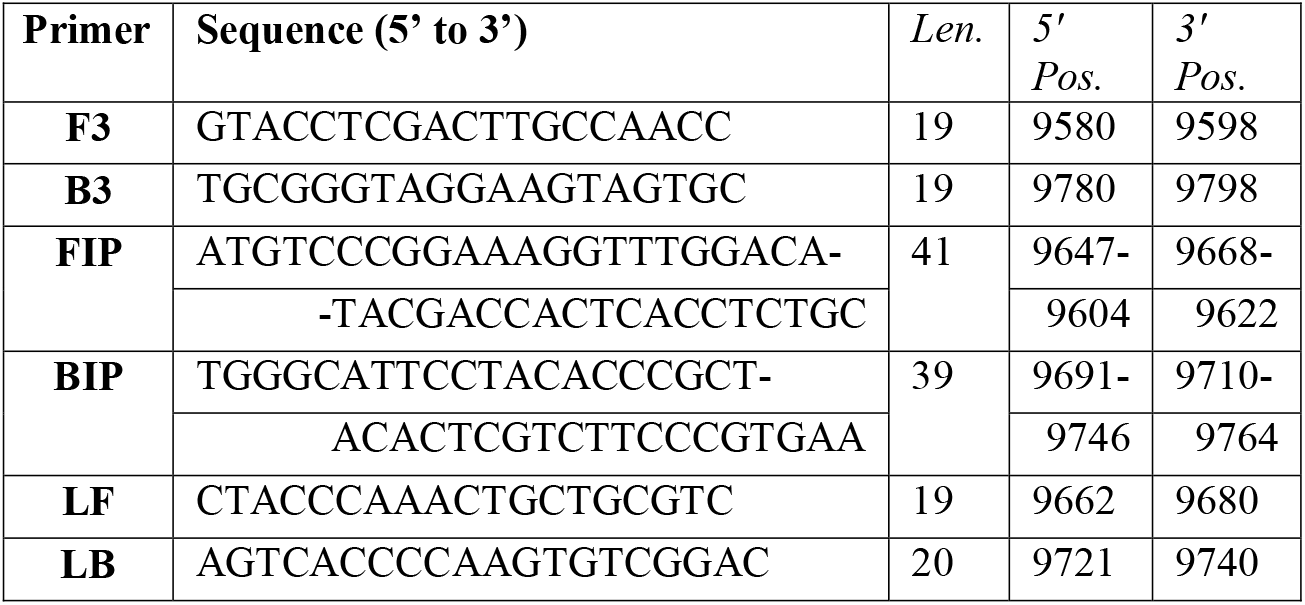
Primer candidate 1 sequences generated using the PrimerExplorerV5 software with respective loop primers (LF and LB).

**Table 2.**
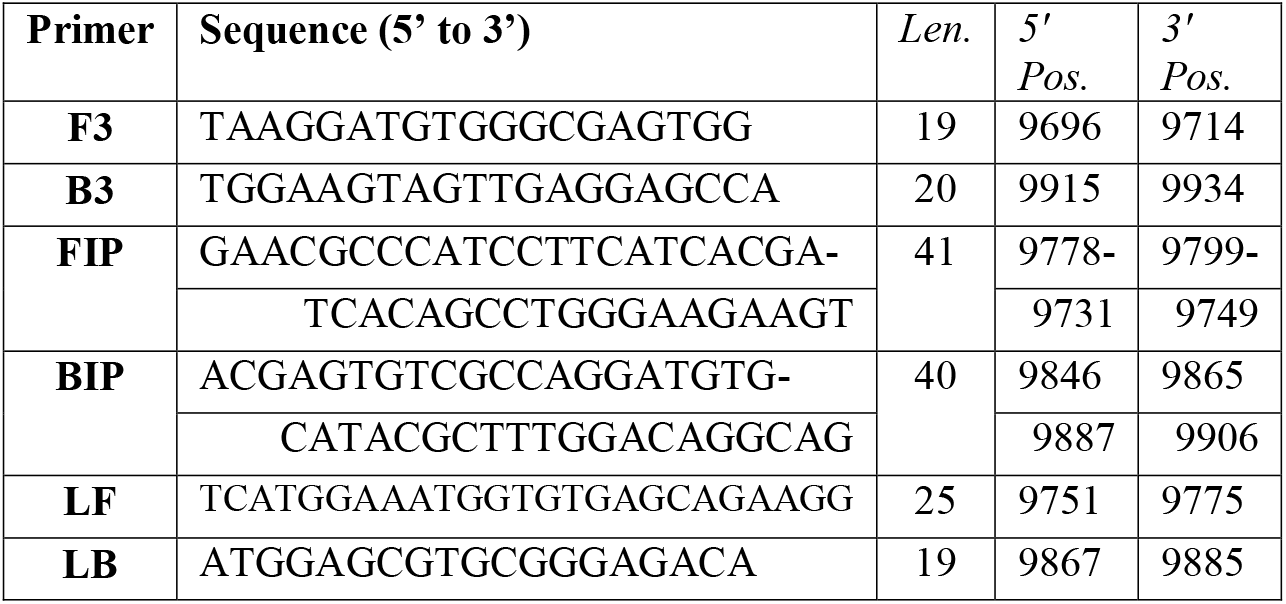
Primer candidate 2 sequences generated using the PrimerExplorerV5 software with respective loop primers (LF and LB).

LAMP primers were determined using the PrimerExplorerV5 software and the AF311056.1 DTV accession (Kuno et al., 2001). The following primers have been designed using the PrimerExplorerV5 software and have been experimentally tested in the laboratory. Primer set candidates 1 and 2 are displayed with length and position on accession sequence in Tables 1 and 2, respectively. Four primers were designed within the first iteration of software for binding to a target sequence. Figure 1 panel A depicts the amplification process that occurs to create the loop products generated by *Bst* polymerase. The inner primers denoted Forward Inner Primer (FIP) and Backward Inner Primer (BIP) are the central component of amplification and loop structure formation. Two additional primers F3 and B3, complementary to F3c and B3c respectively, can bind to the target DNA and displace the amplification product. This allows a repetitive, rapid amplification process. The final two primers Loop Front (LF) and Loop Back (LB) are not pictured in Figure 1 but are able to further amplify and accelerate the loop creation process by providing templates. The reaction is still possible without LF and LB, but a greater limit of detection can be achieved using loop primers. The constructs for LF and LB were generated by inserting the initial primer file into the PrimerExplorerV5 software and following the same protocol.

**Figure 1.**
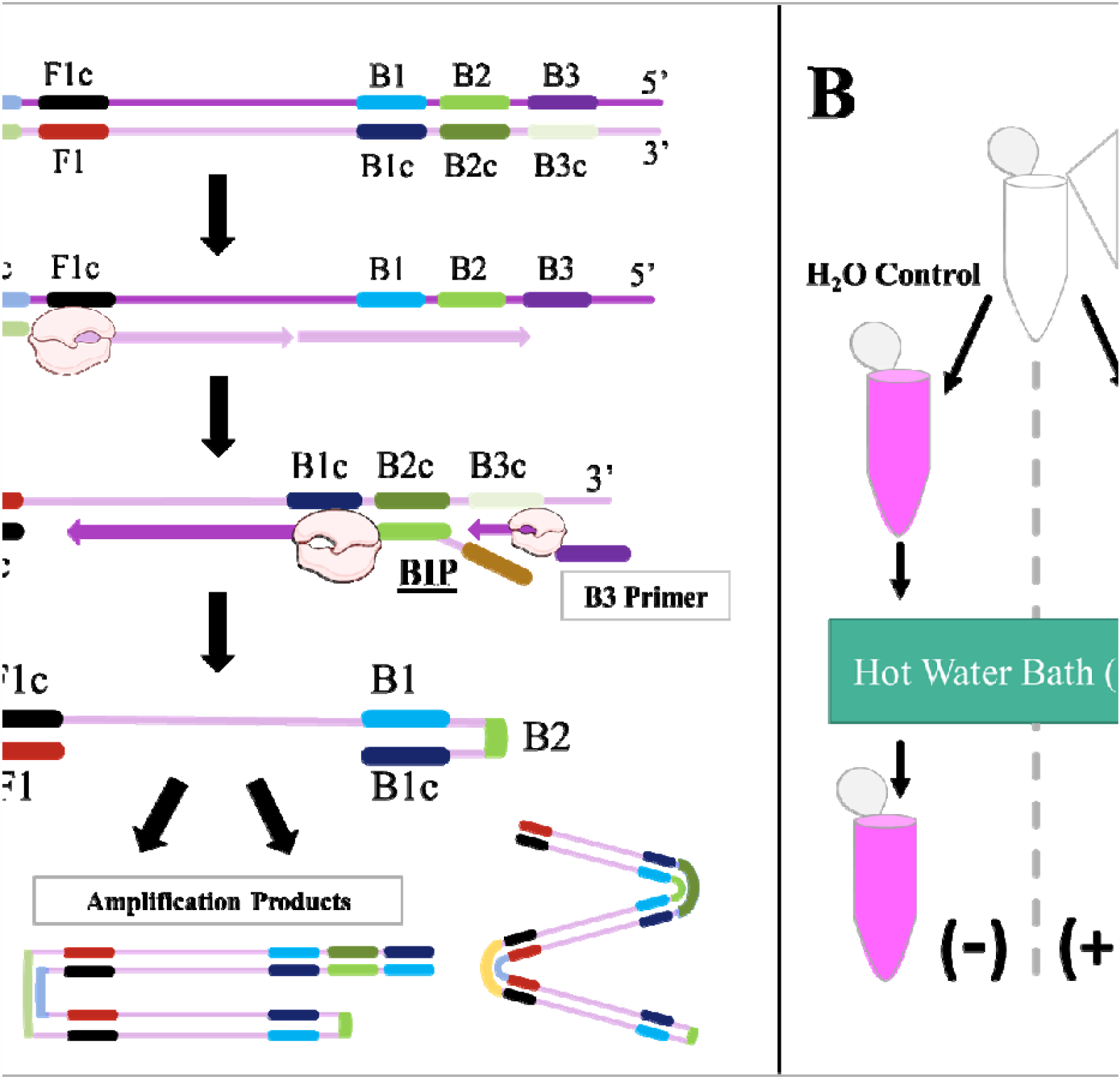
**(A) General schematic of LAMP proliferation into ladder structures.** Each vertical black arrow indicates a new step of the process for amplification. Three prime and five prime ends are indicated on each side of the strands of DNA as well as the LAMP products. Pink and purple horizontally positioned arrows indicate polymerase activity and are shown in conjunction with the polymerase structure. The initial double stranded DNA is representative of the target DNA cloned into a plasmid. F1, F2, and F3 represent the forward primer sequences and are contained within the FIP, F3, and LF primers. F1c, F2c, and F3c represent coding sequences of target DNA. This same principle applies to the B1, B2, B3, B1c, B2c, B3c, BIP and LB sequences in a similar manner to create the end loop product of LAMP. Continual amplification events result in variable ladder structures as depicted. **(B) General reaction scheme for a colorimetric LAMP assay**. Reagents are listed outside first reaction vessel (top) and diverge based on using either 1 μL of experimental sample or water control. The results from this procedure include the stagnant pink color for the negative control, and either a color change to yellow, indicating a positive result, or remain pink indicating a negative result for the experimental DNA.

### Loop-Mediated Isothermal Amplification (LAMP)

For the purposes of this experimentation, the optimal temperature for primer candidate 1 was determined to be 59.3°C and the optimal time was twenty minutes. The Colorimetric Warmstart® MasterMix (New England Biolabs) contains nucleotides, *Bst* polymerase, 16.0 mM Magnesium Sulfate, and pH sensitive indicator phenol red. The general reaction scheme for colorimetric based LAMP assays is seen in Figure 1 panel B. After the addition of genetic material and molecular grade water to Warmstart® (Table 3), the reaction will proceed in either a hot water bath or a thermocycler held at constant temperature. Results are visualized by the naked eye to be pink (negative) or yellow (positive).

**Table 3.** Reagents added to reaction vessels to perform the colorimetric LAMP assay based on New England Biolabs® protocol.

|  | *Experimental* | *Control* |
| --- | --- | --- |
| Warmstart® Colorimetric LAMP 2X Master Mix | 12.5 µL | 12.5 µL |
| LAMP Primer Mix 10X | 2.5 µL | 2.5 µL |
| Target Genetic Sample | 1 µL | N/A |
| Molecular Grade H<sub>2</sub>O | 9 µL | 10 µL |

### Transformation and purification of plasmid DNA

A plasmid was created (GenScript) containing a 500 bp synthetic region of DTV’s RdRp, with flanking *EcoRI* and *XhoI* restriction enzyme sites. The plasmid was transformed into DH5α chemically competent *E. coli* for 2 minutes at 37°C and plated on nutrient agar plates containing ampicillin. DNA stocks were harvested from single clones by plasmid miniprep kits (Qiagen). The desired cDNA was digested from the vector by EcoRI and XhoI, run on an agarose gel, and purified from the gel with a gel extraction kit (Qiagen). The cDNA from this purification process was used for the experimentation as the DTV RdRp coding strand sample.

## Discussion

From the analysis methods, we concluded that these sequences are statistically significant (*p* < 0.05) to move forward with the RdRp targeted primer design. A 2,000 base-pair sequence was chosen from the DTV NS5a coding region GenBank accession AF311056.1, which includes the coding region for the RdRp. The codon-based test resulted in rejecting the null hypothesis for comparing the four different accessions, with *p* < 0.05 for each entry (Table 4). These four accessions represent four divergent strains within the United States and contain great nucleotide conservation. An evolutionary analysis showed the branching of 22 accessions from GenBank including complete genomes and independent RdRp sequences and is visualized in Figure 2. Alignment was performed against the full genome to match the RdRp sequences. The maximum divergence was minimal across all accessions, and primer development was focused in an appropriate area of the DTV genome.

**Table 4.** Codon-based Test of Neutrality for DTV sequences found in the United States.*

|  | **EU338403.1** | **MG196295.1** | **NC003687.1** | **HM991145.1** |
| --- | --- | --- | --- | --- |
| **EU338403.1** | --- | -2.5830 | -3.7536 | 4.4586 |
| **MG196295.1** | *0.0110* | --- | -15.7194 | -3.2327 |
| **NC003687.1** | *0.0003* | *0.0000* | --- | -3.2986 |
| **HM991145.1** | *0.0000* | *0.0016* | *0.0013* | --- |

\*The probability of rejecting the null hypothesis of strict-neutrality ( $dN = dS$ ) (below diagonal) is shown. Values of P less than 0.05 are considered significant at the 5% level and are highlighted. The test statistic ( $dN - dS$ ) is shown above the diagonal. $dS$ and $dN$ are the numbers of synonymous and nonsynonymous substitutions per site, respectively. The variance of the difference was computed using the analytical method. Analyses were conducted using the Nei-Gojobori method (Felsenstein J. 1985). This analysis involved 4 nucleotide sequences. All ambiguous positions were removed for each sequence pair (pairwise deletion option). There was a total of 3556 positions in the final dataset. Evolutionary analyses were conducted in MEGA X (Kumar et al. 2018).

**Figure 2.**
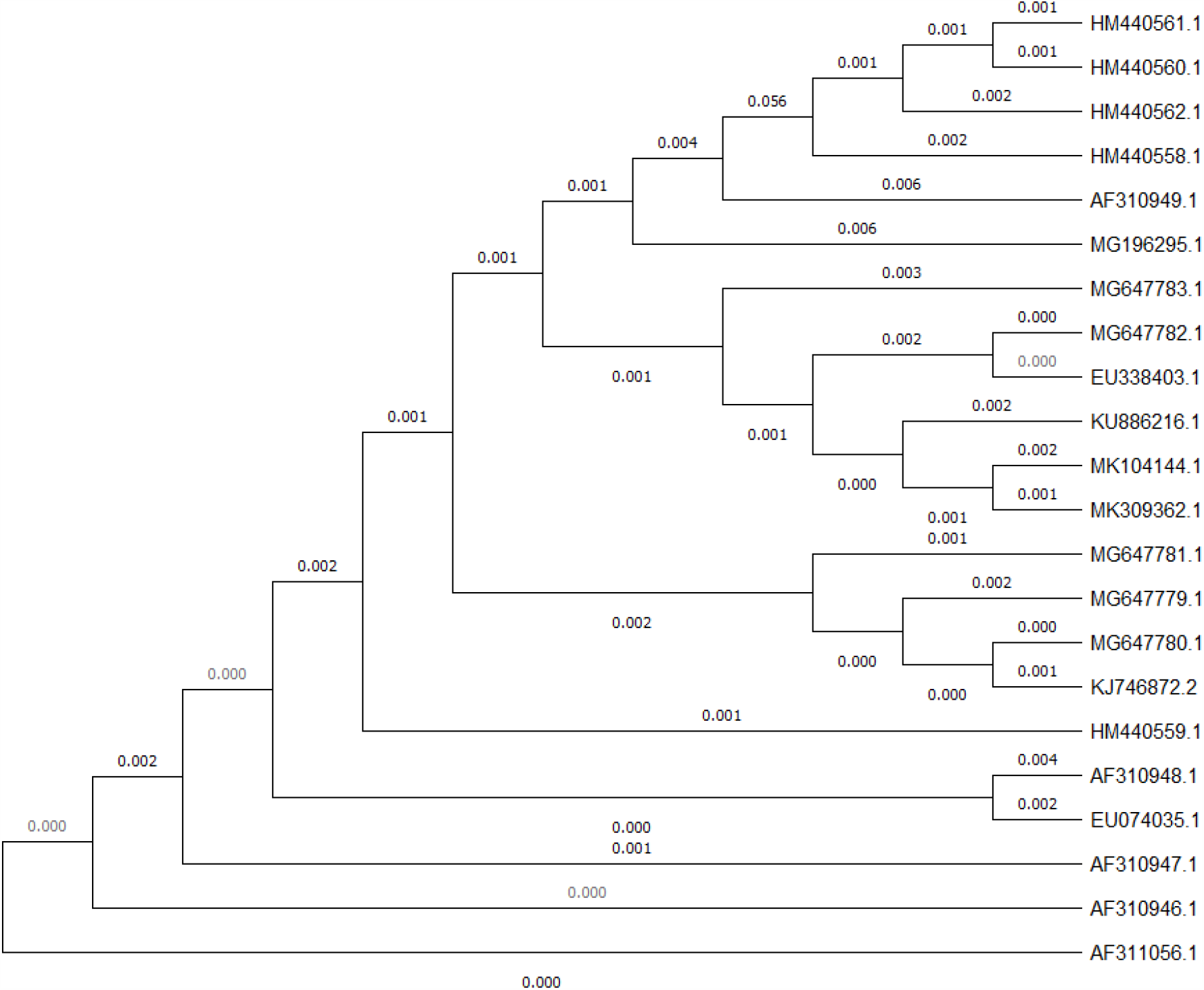
Evolutionary Relationships of Powassan Virus Lineage II, Deer Tick Virus. The evolutionary history was inferred using the Maximum Likelihood method and Tamura-Nei model (Tamura et al. 1993, Felsenstein et al. 1985). The tree with the highest log likelihood (−20885.61) is shown. The tree is drawn to scale, with branch lengths in the same units as those of the evolutionary distances used to infer the phylogenetic tree. Initial tree(s) for the heuristic search were obtained automatically by applying Neighbor-Join and BioNJ algorithms to a matrix of pairwise distances estimated using the Tamura-Nei model, and then selecting the topology with superior log likelihood value (Felsenstein J. 1985). This analysis involved 22 nucleotide sequences and consisted of 1000 bootstrap replications. There was a total of 10843 positions in the final dataset. Evolutionary analyses were conducted in MEGA X (Kumar et al. 2018).

Primer candidate 1 displayed potential and consistency within LAMP assays. Dimerization events were unlikely to occur and provided consistent results. Figure 3 displays typical results from our selected primer sets. When LAMP is run with these primers, a colorimetric change is visualized, and the corresponding products are visualized following electrophoresis on an agarose gel. Controls of nonspecific DNA and water provide negative results as expected. These are desirable traits with LAMP primers to avoid false positives or false negatives. Additional primer sets were designed to confirm our design strategies. The optimal temperature at which this primer candidate function was 59.3°C (Figure 3.D).

**Figure 3.**
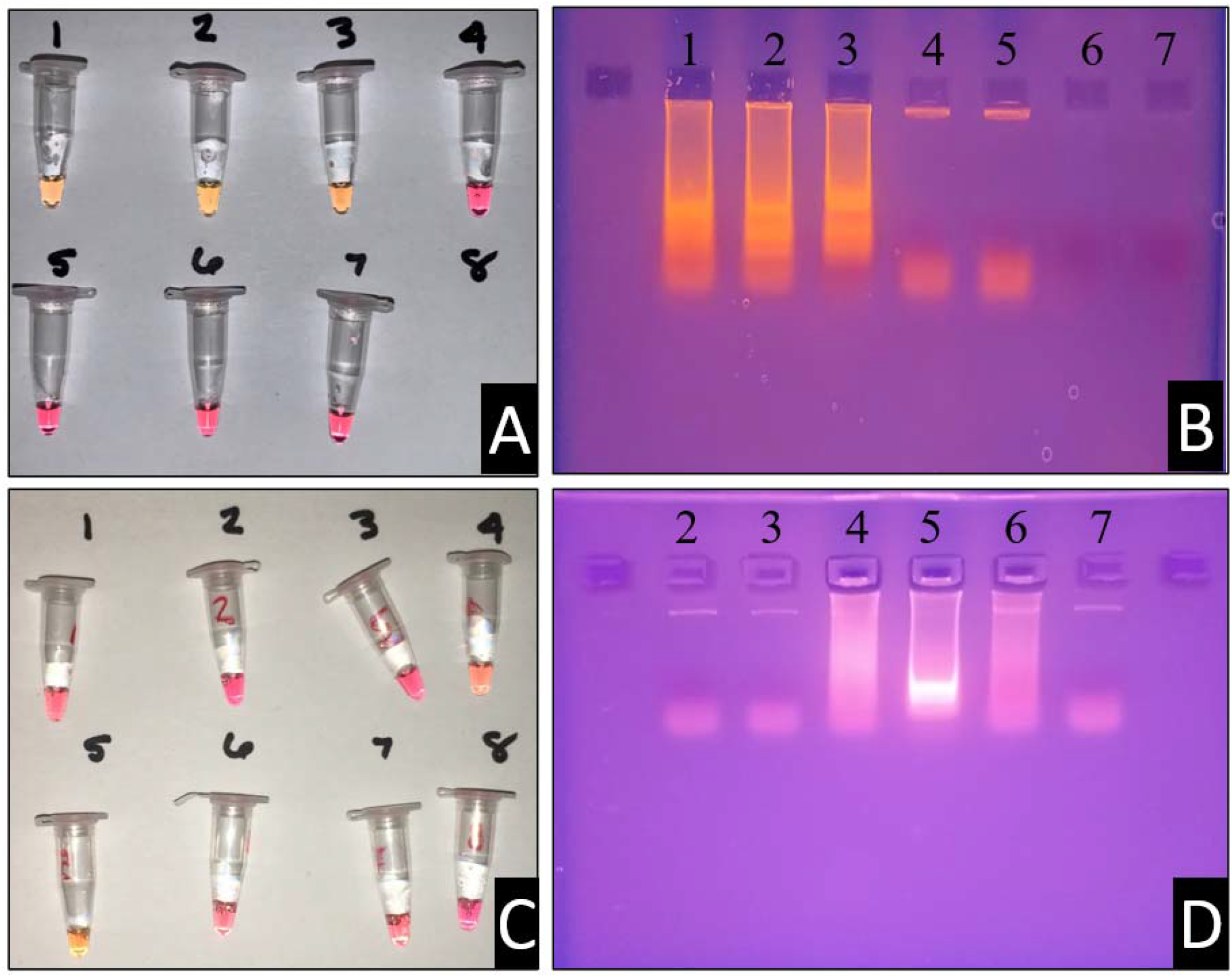
**(A) Colorimetric reaction vessels for primer set 1**.**0 with the following reaction vessel containments:** 1 – Primer w/DNA, 2 – Primer w/DNA, 3 – Primer w/DNA, 4 – Primer w/o DNA, 5 – Primer w/o DNA, 6 – No Primer/DNA, 7 – No Primer/DNA. **(B) Ethidium bromide electrophoresis gel with samples of panel (A)**. The containments of each lane correspond with the reaction vessels as indicated. **(C) Temperature gradient assay for LAMP primer set 1**.**0**. Each of the reaction vessels (left) were also run with negative controls, not pictured. Reaction vessel numbers correspond with the ethidium bromide gel pictured in box (D), with the exception of vessel (1) and (8). From vessel (1) to (8), the temperature gradient follows the order (in °C): 70.8, 69.3, 67.8, 64.8, 59.3, 57.8, 56.3, and 54.8.

The second primer set displayed dimerization concerns after looking closely at the dimerization rate of the 5’ end of the F3 primer. This observation was denoted after consistent false positives were obtained with primer candidate 2. The instability of these primers was directly correlated to the instability of the F3 primer being more positive than dG = −4.00 (Table 2). This observation can be regarded as a learning guide for future primer design experimentation and allowed for a consistency check with the first iteration generated.

RNA was generated from cDNA using a HiScribe T7 Quick High Yield RNA Synthesis Kit (New England Biolabs #E2050S) and resulted in similar amplification results as discussed (data not shown). With some variability of these primer sets, it would be better to investigate additional primer candidates before comparing in RNA-doped tick lyase or serum samples. LAMP technology has shown effective detection for both DNA and RNA material and would be simple to implement even for ssRNA viruses like DTV in *flaviviridae*. RT-LAMP includes minimal adaptation to implement in coordination with an additional reagent.

Future primer sets will be compared to these initial two primer candidates and put through a comparative analysis for accuracy and limit of detection. An additional primer candidate has already been synthesized and experimentation shows additional promise compared to candidate 1. The efforts of this research are to bring attention to this pathosystem and the lack of robust diagnostic testing for downstream health effects. DTV is a recent pathogen of interest and needs additional detection methods to monitor the long-term effects and spread. In the creation of a LAMP detection system point-of-care and field diagnostic, additional tools can be enacted to understand the spread of this virus. Primer set 1 was an excellent candidate for the detection of *in vitro* genetic material of the virus. Our second primer set was faulty but further improved primer design hypotheses and tactics. Further validation is currently being investigated for newer primer sets. Additional experimentation will be performed with RNA expression samples, tick samples spiked with DNA and RNA, and additional collaborations with state environmental agencies for tick testing. In addition to these tests, further experiments should be performed on isolates from varying regions in conjunction with further bioinformatic approaches. Through these efforts we hope to further promote mitigation tactics and reduce the number of infections in North America and beyond.

## Data Availability

No external data was generated/is required outside of this manuscript.

## Acknowledgments

We would like to thank the Lebanon Valley College Biology Department for financially supporting this research, as well as the Beta Beta Beta Biological Honors Society National Research Grant for the 2019-2020 grant received for this project.

## Conflict of Interests

The authors hereby state no conflict of interests.

## Notes

### Competing Interest Statement

The authors have declared no competing interest.

### Author Declarations

Not applicable to this study

### Summary of Updates

Considerations have been taken to improve the manuscript. Figure 1 and Figure 2 have been combined into a single figure. The phylogenetic tree has been updated to include 22 accessions to display the nucleotide conservation.

